# Benzodiazepine Initiation Effect on Mortality Among Medicare Beneficiaries Post Acute Ischemic Stroke

**DOI:** 10.1101/2024.08.18.24312199

**Authors:** Madhav Sankaranarayanan, Maria A. Donahue, Shuo Sun, Julianne D. Brooks, Lee H. Schwamm, Joseph P. Newhouse, John Hsu, Deborah Blacker, Sebastien Haneuse, Lidia M.V.R. Moura

**Author notes:** Corresponding author: Madhav Sankaranarayanan, 321 Franklin St. #2, Cambridge, MA 02139. These authors contributed equally (Sebastien Haneuse, Lidia M.V.R. Moura).

## Abstract

**Rationale:** Despite guideline warnings, older acute ischemic stroke (AIS) survivors still receive benzodiazepines (BZD) for agitation, insomnia, and anxiety despite being linked to severe adverse effects, such as excessive somnolence and respiratory depression. Due to polypharmacy, drug metabolism, comorbidities, and complications during the sub-acute post-stroke period, older adults are more susceptible to these adverse effects. We examined the impact of receiving BZDs within 30 days post-discharge on survival among older Medicare beneficiaries after an AIS.

**Methods:** Using the Medicare Provider Analysis and Review (MedPAR) dataset, Traditional fee-for-service Medicare (TM) claims, and Part D Prescription Drug Event data, we analyzed a random 20% sample of TM beneficiaries aged 66 years or older who were hospitalized for AIS between July 1, 2016, and December 31, 2019. Eligible beneficiaries were enrolled in Traditional Medicare Parts A, B, and D for at least 12 months before admission. We excluded beneficiaries who were prescribed a BZD within 90 days before hospitalization, passed away during their hospital stay, left against medical advice, or were discharged to institutional post-acute care. Our primary exposure was BZD initiation within 30 days post-discharge, and the primary outcome was 90-day mortality risk differences (RD) from discharge. We followed a trial emulation process involving cloning, weighting, and censoring, plus we used inverse-probability-of-censoring weighting to address confounding.

**Results:** In a sample of 47,421 beneficiaries, 826 (1.74%) initiated BZD within 30 days after discharge from stroke admission or before readmission, whichever occurred first, and 6,392 (13.48%) died within 90 days. Our study sample had a median age of 79, with an inter-quartile range (IQR) of 12, 55.3% female, 82.9% White, 10.1% Black, 1.7% Hispanic, 2.2% Asian, 0.4% American Native, 1.5% Other and 1.1% Unknown. After standardization based on age, sex, race/ethnicity, length of stay in inpatient, and baseline dementia, the estimated 90-day mortality risk was 159 events per 1,000 (95% CI: 155, 166) for the BZD initiation strategy and 133 events per 1,000 (95% CI: 132, 135) for the non-initiation strategy, with an RD of 26 events per 1,000 (95% CI: 22, 33). Subgroup analyses showed RDs of 0 events per 1,000 (95% CI: -4, 11) for patients aged 66-70, 3 events per 1,000 (95% CI: -1, 13) for patients aged 71-75, 10 events per 1,000 (95% CI: 3, 23) for patients aged 76-80, 27 events per 1,000 (95% CI: 21, 46) for patients aged 81-85, and 84 events per 1,000 (95% CI: 73, 106) for patients aged 86 years or older. RDs were 34 events per 1,000 (95% CI: 26, 48) and 20 events per 1,000 (95% CI: 11, 33) for males and females, respectively. RDs were 87 events per 1,000 (95% CI: 63, 112) for patients with baseline dementia and 18 events per 1,000 (95% CI: 13, 21) for patients without baseline dementia.

**Conclusion:** Initiating BZDs within 30 days post-AIS discharge significantly increased the 90-day mortality risk among Medicare beneficiaries aged 76 and older and for those with baseline dementia. These findings underscore the heightened vulnerability of older adults, especially those with cognitive impairment, to the adverse effects of BZDs.

## INTRODUCTION

Every year in the United States, about 800,000 individuals have a stroke,^1^ and three-quarters of all strokes occur in adults aged 65 and older, leading to around 610,000 hospital admissions annually.^2^ Acute ischemic stroke (AIS) survivors are often given benzodiazepines (BZDs) during the immediate acute recovery period for periprocedural sedation or to manage insomnia, anxiety, and agitation, despite guideline warnings.^3,4^

BZDs are associated with severe adverse effects, such as cognitive impairment, psychomotor difficulties, excessive somnolence, pneumonia, respiratory depression requiring intubation, falls, and death.^5–11^ Older adults are especially susceptible due to age-related changes in drug metabolism, polypharmacy, and comorbidities.^12^ Patients who experience an AIS are particularly vulnerable to the risks associated with BZD use,^10,13^ with pre-existing brain damage and gait comorbidities compounding the established risks of falls and fall-related injuries. Alarmingly, various studies have shown that up to 15% of stroke patients receive BZDs, often indefinitely, and 50-80% of older patients using BZDs chronically experience adverse effects.^3,6–9,14,15^

Despite state policies and national guidelines to curb BZD prescriptions, usage remains high.^4,14^ There is a lack of data and studies on older adult outcomes among those receiving BZD prescriptions in the sub-acute AIS recovery period. Trials among older adults are infeasible for several reasons: exclusion criteria (e.g., comorbidities, polypharmacy, and functional limitations), recruitment barriers (multimorbidity, disabilities, frailty, etc.), cognitive impairment, consent issues, drug interactions, adverse effects, and high drop-out rates, among others.^16^ We examined the impact of new outpatient BZD prescriptions dispensed 30 days post-AIS discharge among Medicare beneficiaries 65 years or older.

## METHODS

This study followed the Strengthening the Reporting of Observational Studies in Epidemiology (STROBE)^17^ reporting guidelines and was approved by the Mass General Brigham Institutional Review Board. Because we conducted a secondary analysis of data collected for routine care and billing purposes, the requirement for informed consent was waived. The data supporting this study’s findings are collected by The Centers for Medicare & Medicaid Services (CMS) and were made available by CMS with no direct identifiers. All results were aggregated following CMS Cell Suppression Policies. Restrictions apply to the availability of these data, which were used under license for this study. Medicare data are available through CMS with their permission. The code that produced the findings is available upon reasonable request from any qualified investigator.

### Eligibility Criteria

To properly define our target trial, we must establish the criteria for selecting patients in a 20% random sample of Traditional Medicare Claims eligible for said trial. We identified 92,852 patients who were admitted for AIS between July 1, 2016, and December 31, 2019, and enrolled in Traditional Fee-For-Service Medicare with at least one year of enrollment in Parts A (hospital insurance information), B (preventative and medically necessary services or medical insurance information), and D (prescription drug coverage information) before AIS hospitalization. We included patients admitted to an acute hospital and discharged home (short-stay hospitalization). We included patients aged 66 and above in the analysis. We excluded patients with a recorded AIS diagnosis 12 months before hospitalization and patients with one or more recorded outpatient BZD prescriptions within three months before admission. The final eligible sample consisted of 47,421 patients (Figure 1).

**Figure 1.**
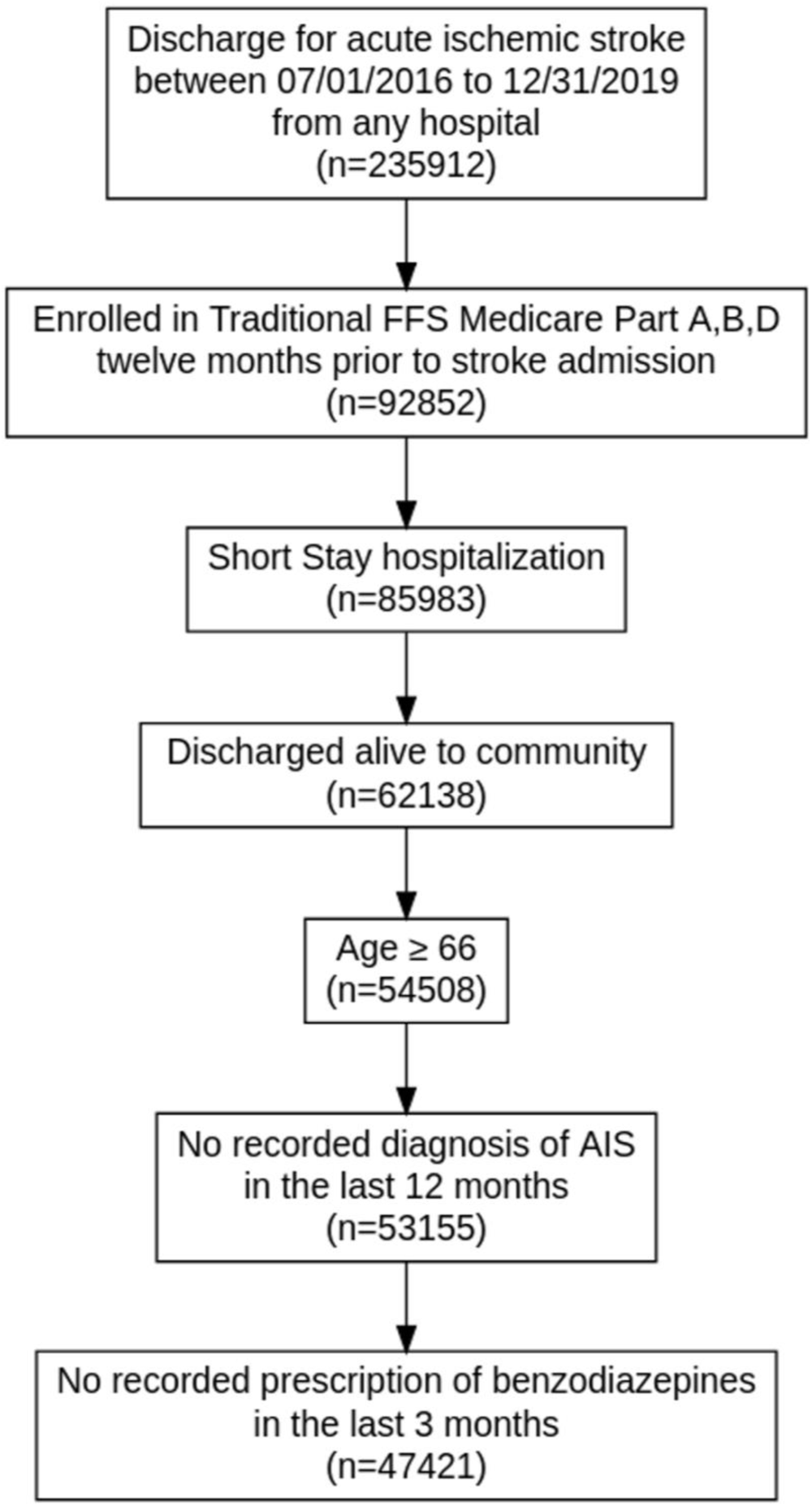
Schematic Describing Eligibility Criteria Describes the sampling process resulting in a sample of 47,421 patients. Medicare data files used: MedPAR (Inpatient data), MBSF (Summary data); Diagnosed for Acute Ischemic Stroke based on ICD-10 codes (I63 and I63.9); Part A: Hospital Insurance, Part B: Medical Insurance; Part D: Drug coverage. AIS, Acute Ischemic Stroke; FFS, Fee-for-Service.

### Study Design

The target trial would randomly assign eligible patients discharged alive after an AIS hospitalization to one of two arms: i) BZD treatment upon discharge and ii) usual care with no BZDs for 30 days post-discharge. We modeled an emulated trial based on this target trial, which involved random assignment of eligible patients upon discharge from AIS hospitalization to one of two arms: i) treatment with BZD within the window extending from discharge to 30 days post-discharge (or readmission date if within the 30 days); ii) no BZDs given within the defined window from discharge to 30 days post-discharge or readmission, whichever occurred first. We assessed the primary outcome of mortality during a follow-up period spanning 90 days starting from discharge. Patients were followed from discharge to the earliest of the following events: mortality, readmission, or end of study (i.e., 90 days post-discharge, including the discharge day). Further details regarding the observational analysis proposed for this emulated target trial can be found in Table 1.

**Table 1.** Description of a Target Trial and The Corresponding Observational Study.

| Target Trial | Emulated trial |
| --- | --- |
| **Eligibility Criteria** |  |
| Discharge for cerebrovascular accident between 07/01/2016 and 12/31/2019 from any hospital, and enrolled in Traditional Medicare Part A, B, D 12 months before stroke admission | Same |
| Age $\geq 66$ | Same |
| Confirmed AIS | Same |
| Admitted to a short-stay hospital | Same |
| No previous history of AIS in the last 12 months before admission | No recorded diagnosis of AIS in the last 12 months before admission |
| No use of BZDs in the last 3 months prior to admission | No recorded prescription of BZDs in the last 3 months before admission |
| **Treatment Strategies** |  |
| Treatment arm: Initiate BZD upon discharge, follow prescription strategy for 30 days<br>Control arm: Subject to usual care without BZDs for 30 days post-discharge | Treatment arm: Initiate BZD within a defined window extending from AIS discharge to either the 30th day post-discharge or readmission date, whichever occurs first.<br>Control arm: Refrain from initiating BZD within a defined window extending from AIS discharge to either the 30th day post-discharge or readmission date, whichever occurs first. |
| **Assignment Procedures** |  |
| Randomized treatment assignment | Emulated randomization* |
| **Outcomes** |  |
| Time to death within 90 days post-discharge | Same |
| **Follow-up** |  |
| Starts at randomization (discharge)<br>Ends at date of all-cause re-admission, death, loss to follow-up or end of study (90 days post-discharge); whichever occurs first | Starts at discharge<br>Ends at date of all-cause re-admission, death (as recorded in Medicare data<sup>†</sup>), loss to follow-up (date of enrollment discontinuation<sup>‡</sup>) or end of study (90 days post-discharge); whichever occurs first |
| **Causal Contrast** |  |
| Intention-to-treat effect | Observational analog of intention-to-treat effect<sup>§</sup> |
| **Statistical Analysis** |  |
| Intention-to-treat effect analysis of time to mortality, accounting for censoring. | Intention-to-treat effect analysis of time to mortality, accounting for censoring and baseline confounding. |

### Individual Characteristics

For each beneficiary, we captured demographic characteristics, i.e., age, documented sex, reported race/ethnicity, dual Medicare and Medicaid eligibility status, and the original reason for Medicare entitlement provided in Medicare’s Master Beneficiary Summary (MBSF) File. We used the self-reported race/ethnicity variable provided in MBSF (White, Black, Hispanic, Asian, American Native, Other, Unknown).^18^

We used acute hospitalization records from the Medicare Provider Analysis and Review (MedPAR) file and outpatient diagnosis codes in the Outpatient and Carrier Claims files. AIS hospitalizations were selected based on AIS ICD-10 code I63.^19^ We took the first hospitalization chronologically for beneficiaries with more than one AIS hospitalization. We defined readmission as any inpatient stay recorded post-stroke in the MedPAR file. We also obtained baseline stroke severity using the National Institutes of Health Stroke Scale (NIHSS) score.^20^ We identified NIHSS scores in the MedPAR files using ICD-10 codes R297.00 to R297.42, and we used the maximum NIHSS score in the case of multiple scores. NIHSS scores range from 0 to 42, with higher values indicating a more severe stroke. We grouped NIHSS scores into two categories: Minor (≤4) and Moderate-to-High (>4).^21^ As an additional measure of stroke severity at baseline, we used the length of stay (LOS) during AIS admission. A shorter LOS has been associated with increased risks of early readmissions and post-discharge mortality.^22^ A U-shaped relationship between LOS and 30-day mortality has been described, where both very short (1-2 days) and long (9-14 days) LOS were associated with high mortality rates among patients with heart failure.^23^

We captured comorbid conditions by leveraging all diagnoses present in the 12 months before stroke admission. For each beneficiary, we evaluated the presence of a pre-admission history of myocardial infarction (MI), congestive heart failure (CHF), peripheral vascular disease (PVD), cardiovascular disease (CVD), chronic obstructive pulmonary disease (COPD), paralysis, diabetes, renal disease, liver disease, rheumatism, and dementia^24^ using ICD-10 diagnosis codes listed in Supplemental Table S1 and the National Cancer Institute’s SEER-Medicare: Comorbidity SAS Macros.^42,43^ This algorithm requires that for outpatient claims, a patient’s diagnoses (not just specific codes) must appear on at least two different claims more than 30 days apart. Additionally, we have death dates from the master beneficiary summary file (MBSF). We describe the characteristics of eligible patients stratified by BZD initiation strategy in Table 2.

**Table 2.**
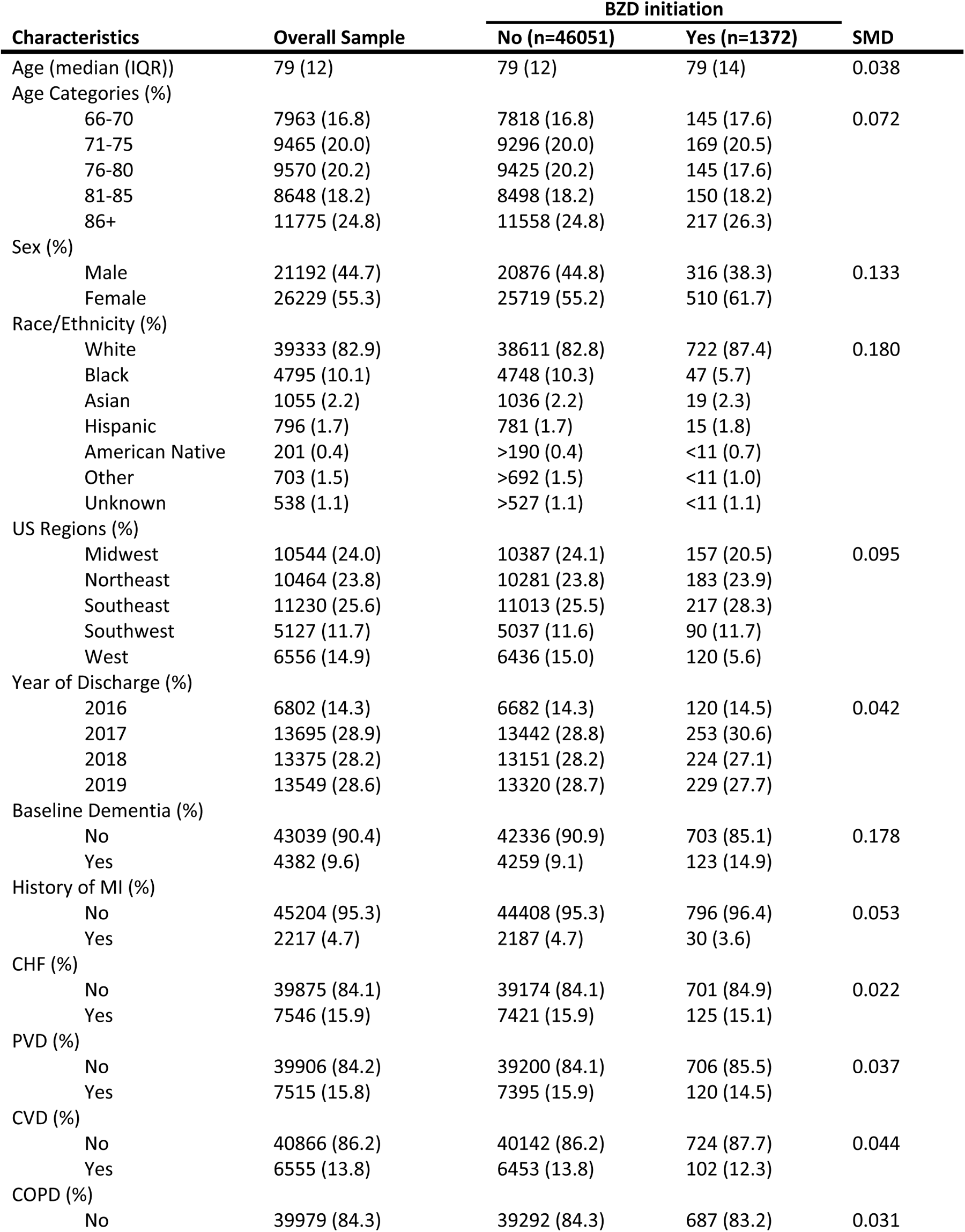
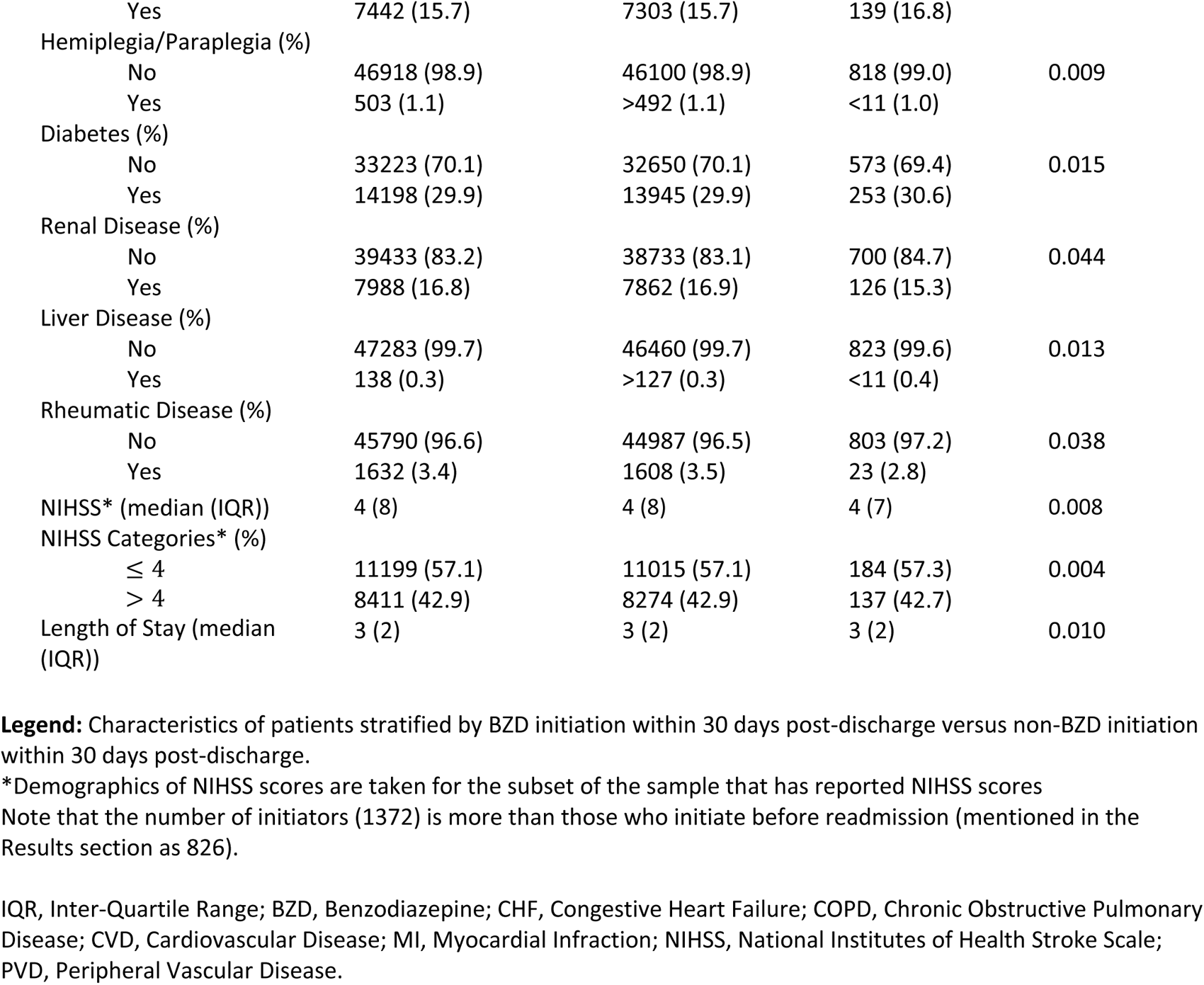
Characteristics of Patients Stratified by Benzodiazepine Initiation.

| Characteristics | Overall Sample | BZD initiation |  | SMD |
| --- | --- | --- | --- | --- |
|  |  | No (n=46051) | Yes (n=1372) |  |
| Age (median (IQR)) | 79 (12) | 79 (12) | 79 (14) | 0.038 |
| Age Categories (%) |  |  |  |  |
| 66-70 | 7963 (16.8) | 7818 (16.8) | 145 (17.6) | 0.072 |
| 71-75 | 9465 (20.0) | 9296 (20.0) | 169 (20.5) |  |
| 76-80 | 9570 (20.2) | 9425 (20.2) | 145 (17.6) |  |
| 81-85 | 8648 (18.2) | 8498 (18.2) | 150 (18.2) |  |
| 86+ | 11775 (24.8) | 11558 (24.8) | 217 (26.3) |  |
| Sex (%) |  |  |  |  |
| Male | 21192 (44.7) | 20876 (44.8) | 316 (38.3) | 0.133 |
| Female | 26229 (55.3) | 25719 (55.2) | 510 (61.7) |  |
| Race/Ethnicity (%) |  |  |  |  |
| White | 39333 (82.9) | 38611 (82.8) | 722 (87.4) | 0.180 |
| Black | 4795 (10.1) | 4748 (10.3) | 47 (5.7) |  |
| Asian | 1055 (2.2) | 1036 (2.2) | 19 (2.3) |  |
| Hispanic | 796 (1.7) | 781 (1.7) | 15 (1.8) |  |
| American Native | 201 (0.4) | >190 (0.4) | <11 (0.7) |  |
| Other | 703 (1.5) | >692 (1.5) | <11 (1.0) |  |
| Unknown | 538 (1.1) | >527 (1.1) | <11 (1.1) |  |
| US Regions (%) |  |  |  |  |
| Midwest | 10544 (24.0) | 10387 (24.1) | 157 (20.5) | 0.095 |
| Northeast | 10464 (23.8) | 10281 (23.8) | 183 (23.9) |  |
| Southeast | 11230 (25.6) | 11013 (25.5) | 217 (28.3) |  |
| Southwest | 5127 (11.7) | 5037 (11.6) | 90 (11.7) |  |
| West | 6556 (14.9) | 6436 (15.0) | 120 (5.6) |  |
| Year of Discharge (%) |  |  |  |  |
| 2016 | 6802 (14.3) | 6682 (14.3) | 120 (14.5) | 0.042 |
| 2017 | 13695 (28.9) | 13442 (28.8) | 253 (30.6) |  |
| 2018 | 13375 (28.2) | 13151 (28.2) | 224 (27.1) |  |
| 2019 | 13549 (28.6) | 13320 (28.7) | 229 (27.7) |  |
| Baseline Dementia (%) |  |  |  |  |
| No | 43039 (90.4) | 42336 (90.9) | 703 (85.1) | 0.178 |
| Yes | 4382 (9.6) | 4259 (9.1) | 123 (14.9) |  |
| History of MI (%) |  |  |  |  |
| No | 45204 (95.3) | 44408 (95.3) | 796 (96.4) | 0.053 |
| Yes | 2217 (4.7) | 2187 (4.7) | 30 (3.6) |  |
| CHF (%) |  |  |  |  |
| No | 39875 (84.1) | 39174 (84.1) | 701 (84.9) | 0.022 |
| Yes | 7546 (15.9) | 7421 (15.9) | 125 (15.1) |  |
| PVD (%) |  |  |  |  |
| No | 39906 (84.2) | 39200 (84.1) | 706 (85.5) | 0.037 |
| Yes | 7515 (15.8) | 7395 (15.9) | 120 (14.5) |  |
| CVD (%) |  |  |  |  |
| No | 40866 (86.2) | 40142 (86.2) | 724 (87.7) | 0.044 |
| Yes | 6555 (13.8) | 6453 (13.8) | 102 (12.3) |  |
| COPD (%) |  |  |  |  |
| No | 39979 (84.3) | 39292 (84.3) | 687 (83.2) | 0.031 |
| Yes | 7442 (15.7) | 7303 (15.7) | 139 (16.8) |  |
| Hemiplegia/Paraplegia (%) |  |  |  |  |
| No | 46918 (98.9) | 46100 (98.9) | 818 (99.0) | 0.009 |
| Yes | 503 (1.1) | >492 (1.1) | <11 (1.0) |  |
| Diabetes (%) |  |  |  |  |
| No | 33223 (70.1) | 32650 (70.1) | 573 (69.4) | 0.015 |
| Yes | 14198 (29.9) | 13945 (29.9) | 253 (30.6) |  |
| Renal Disease (%) |  |  |  |  |
| No | 39433 (83.2) | 38733 (83.1) | 700 (84.7) | 0.044 |
| Yes | 7988 (16.8) | 7862 (16.9) | 126 (15.3) |  |
| Liver Disease (%) |  |  |  |  |
| No | 47283 (99.7) | 46460 (99.7) | 823 (99.6) | 0.013 |
| Yes | 138 (0.3) | >127 (0.3) | <11 (0.4) |  |
| Rheumatic Disease (%) |  |  |  |  |
| No | 45790 (96.6) | 44987 (96.5) | 803 (97.2) | 0.038 |
| Yes | 1632 (3.4) | 1608 (3.5) | 23 (2.8) |  |
| NIHSS* (median (IQR)) | 4 (8) | 4 (8) | 4 (7) | 0.008 |
| NIHSS Categories* (%) |  |  |  |  |
| ≤ 4 | 11199 (57.1) | 11015 (57.1) | 184 (57.3) | 0.004 |
| > 4 | 8411 (42.9) | 8274 (42.9) | 137 (42.7) |  |
| Length of Stay (median (IQR)) | 3 (2) | 3 (2) | 3 (2) | 0.010 |

**Legend:** Characteristics of patients stratified by BZD initiation within 30 days post-discharge versus non-BZD initiation within 30 days post-discharge.
\*Demographics of NIHSS scores are taken for the subset of the sample that has reported NIHSS scores
Note that the number of initiators (1372) is more than those who initiate before readmission (mentioned in the Results section as 826).
IQR, Inter-Quartile Range; BZD, Benzodiazepine; CHF, Congestive Heart Failure; COPD, Chronic Obstructive Pulmonary Disease; CVD, Cardiovascular Disease; MI, Myocardial Infarction; NIHSS, National Institutes of Health Stroke Scale; PVD, Peripheral Vascular Disease.

### Benzodiazepine Exposure and Treatment Assignment Procedures

In the target trial emulation, we obtained information on BZD prescriptions from outpatient pharmacy data. The BZD list can be found in Table S1 in the Supplement.

We addressed immortal time and confounding bias by deploying cloning techniques.^25–27^ Every patient is “assigned” to both the control and treatment arm and “observed” until they deviate from the protocol of the given arm. This effectively doubles our sample size, giving us perfectly balanced arms concerning baseline characteristics. We accounted for this artificial doubling of the sample by “artificially” censoring the person-time for each patient clone when the actual treatment deviates from the assigned treatment (e.g., a patient initiated on BZD within the 30-day exposure window would have a clone in the control arm, contributing person-time until their initiation time). With this approach, we aligned the discharge and assignment times directly. We accounted for the selection bias due to this artificial censoring by using the inverse probability of censoring weighting (IPCW) on the observations. We describe the cloning method in Figure S1 and “Technical Details” in the Supplement.

### Statistical Analysis

To evaluate the effect of BZD initiation within the defined post-AIS exposure period on 90-day mortality, we estimated the survival probability of the patients using model-based predictions. After cloning and censoring the data, we weighted the expanded data using stabilized IPCWs to account for artificial censoring. The models for IPCW were pooled linear logistic regressions over person days, including age, sex, race/ethnicity, baseline dementia, and LOS as covariates, along with potential interactions between them, separately for the two treatment strategies. For comparison, we estimated the unadjusted Kaplan-Meier and an unadjusted cubic spline model-based survival probabilities of mortality in the 90 days using the cloned dataset (see Figure S2 in the Supplement).

We fitted a pooled logistic regression model for mortality as a function of the following covariates: treatment strategy, a B-spline function with 5 degrees of freedom of time (measured in days post-discharge), and an interaction term between the treatment strategy and the B-spline for time to allow for time-varying effects using the expanded weighted data. We obtained estimated survival probabilities for each day under each treatment strategy and estimated the 90-day mortality risk difference.

We used the bootstrap method based on 500 re-samplings to obtain 95% Confidence Intervals (CI), accounting for sample size inflation by cloning. See Technical Details in the Supplement for the specifics of our statistical analysis.

### Missing Data

We found no missing demographic information for the patients in our selected cohort. There was minimal missingness in the selected cohort’s baseline medical conditions (4%). We assumed no baseline conditions for those without a recorded one. For NIHSS scores, there was a high missingness (58.6%). We performed a complete case analysis using NIHSS scores, sex, and dementia (please see the Supplement for these analyses).

### Pre-planned Stratified Analysis

We repeated the analyses above, stratifying on age categories since BZDs are viewed as more detrimental to older patients. We also repeated our analyses stratified on baseline dementia, as BZD has more damaging effects in those with dementia.^28^ Lastly, we stratified on sex since females are prescribed these medications more frequently and are more likely to experience side effects due to physiological and pharmacodynamic differences. We also conducted a secondary analysis stratifying on NIHSS scores (<4 vs. >4) in patients with a reported NIHSS score, as BZDs may be more detrimental to patients with more severe AIS.

### Secondary Analysis

In secondary analysis, we sought to include a marker of stroke severity at baseline because it has been associated with prescription probability and mortality risk. We examined the proportion of reported NIHSS scores, which was around 43% for the Medicare sample, and we relegated the use of NIHSS scores as a metric for stroke severity to the Supplement (Table S4, Figure S6-S11, and the Technical Details section).

## RESULTS

### Study Population Characteristics

In a sample of 235,912 beneficiaries discharged for an AIS during the study period, 47,421 patients met our inclusion criteria (Figure 1). Of these, 826 (1.74%) initiated BZDs within 30 days after discharge from the stroke admission (or before readmission, whichever occurred first), and 6,392 (13.48%) passed away within 90 days. The median age of our study sample was 79 (IQR 12). Our sample consisted of 55.3% female beneficiaries, 82.9% White, 10.1% Black/African American, 1.7% Hispanic, 2.2% Asian, and 0.4% North American Native. We provide additional patient demographic/clinical characteristics by BZD initiation strategy in Table 2.

### Outcome: Mortality

After target trial emulation with cloning and adjustment for artificial censoring, the standardized 90-day risk of mortality was 159 events per 1000 (95% CI: 155,166) for those under the BZD initiation strategy and 133 events per 1000 (95% CI: 132,135) for those under the non-initiation strategy, which resulted in a risk difference of 26 events per 1000 patients (95% CI: 22,33). Figure 2 shows the standardized survival functions.

**Figure 2.**
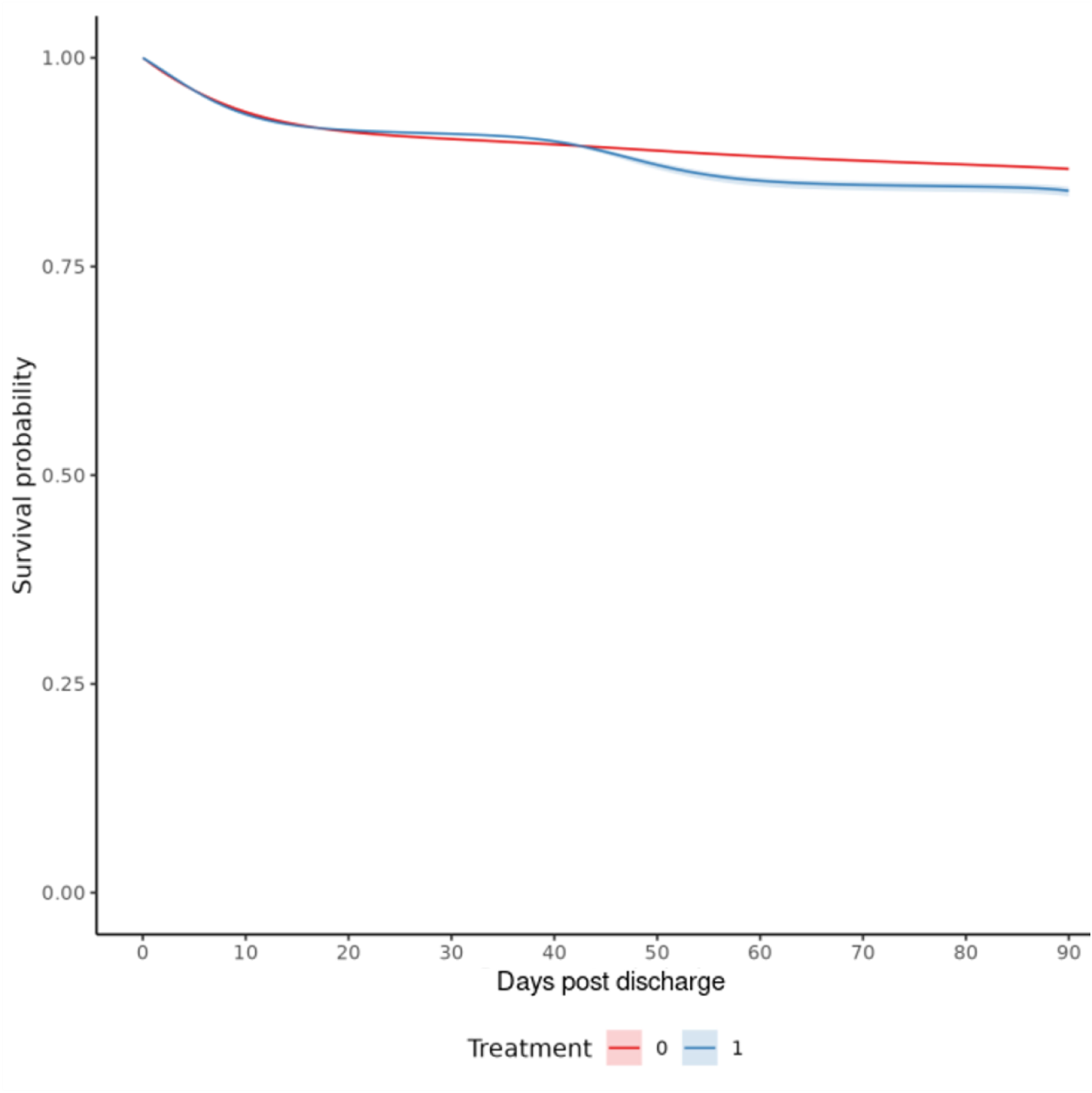
Standardized Survival Curve by Benzodiazepine Initiation Strategy. pooled logistic regression survival curves for all patients. **Blue:** Strategy for benzodiazepine initiation within 30 days post-discharge from AIS admission. **Red:** Strategy for no benzodiazepine initiation within 30 days post-discharge from AIS admission. Shaded areas: 95% CIs were constructed using Bootstrap with 500 replications.

Looking at age categories, the 90-day risk differences were 0 events per 1000 (95% CI: -4, 11) for patients aged 66-70, 3 events per 1000 (95% CI: -1, 13) for patients aged 71-75, 10 events per 1000 (95% CI: 3, 23) for patients aged 76-80, 27 events per 1000 (95% CI: 21, 46) for patients aged 81-85, and 84 events per 1000 (95% CI: 73, 106) for patients aged 86 years or older. Risk differences were 34 events per 1000 (95% CI: 26,48) and 20 events per 1000 (95% CI: 11,33) for male and female patients, respectively. Risk differences were 87 events per 1000 (95% CI: 63,112) for patients with baseline dementia and 18 events per 1000 (95% CI: 13,21) for patients without baseline dementia (Figure 3). The standardized 90-day risks of mortality stratified by age categories, sex, and baseline dementia are displayed in Table 3. The analysis in the subpopulation with reported NIHSS scores yielded results similar to our primary analysis (“Additional Analysis” in the Supplement). The Supplementary Text provides the Statistical Code used for the primary analysis.

**Figure 3.**
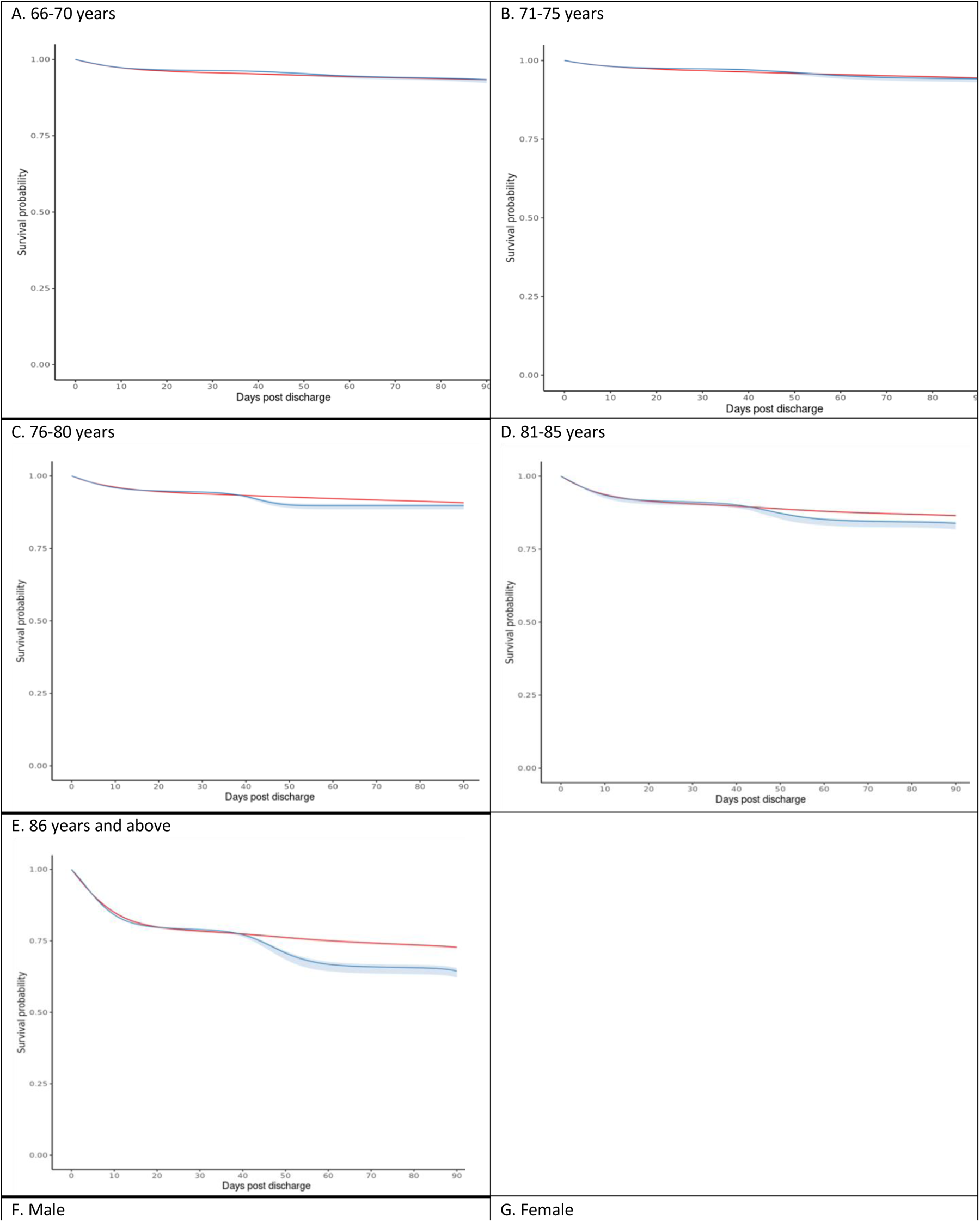
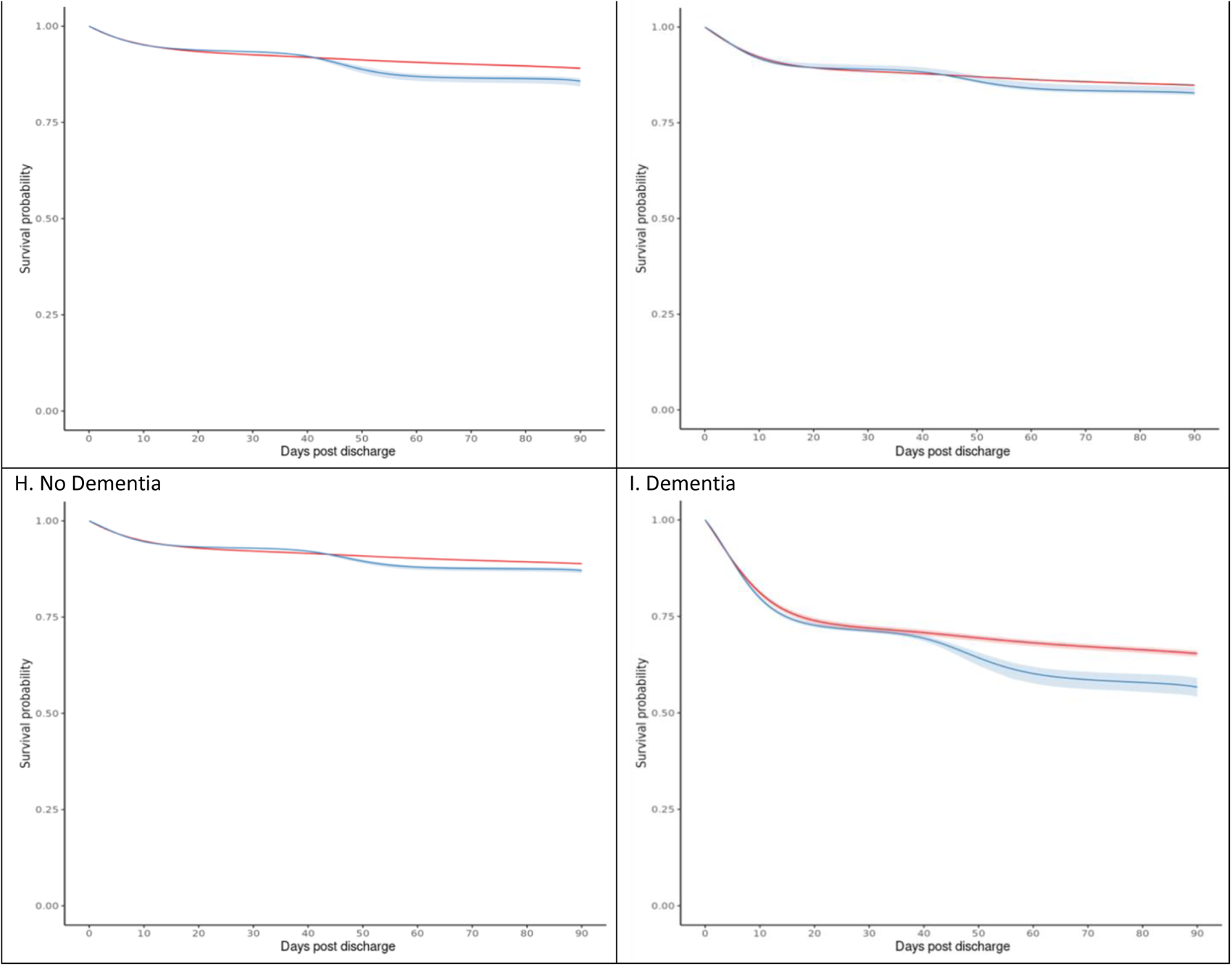
Standardized Survival Curves, Stratified Based on Age, Sex, and Dementia. pooled logistic regression survival curves for patients of the given strata. **Blue:** Strategy for benzodiazepine initiation within 30 days post-discharge. **Red:** Strategy for no benzodiazepine initiation within 30 days post-discharge. Shaded areas: 95% of CIs were constructed using Bootstrap with 500 replications.

**Table 3.** Standardized 90-Day Risks of Mortality.

| Covariate/<br>Stratum | Initiators |  | Non-initiators |  | Risk Difference |  |
| --- | --- | --- | --- | --- | --- | --- |
|  | Estimate | 95% CI | Estimate | 95% CI | Estimate | 95% CI |
| *Overall* | 159 | (155,166) | 133 | (132,135) | 26 | (22,33) |
| **Age** |  |  |  |  |  |  |
| 66 to 70 years | 55 | (50,66) | 55 | (53,58) | 0 | (-4,11) |
| 71 to 75 years | 69 | (66,81) | 66 | (63,69) | 3 | (-1,13) |
| 76 to 80 years | 103 | (96,115) | 92 | (90,95) | 10 | (3,23) |
| 81 to 85 years | 161 | (156,182) | 135 | (131,139) | 27 | (21,46) |
| 86+ | 356 | (345,379) | 272 | (268,277) | 84 | (73,106) |
| **Sex** |  |  |  |  |  |  |
| Male | 143 | (136,157) | 110 | (107,112) | 34 | (26,48) |
| Female | 172 | (165,173) | 152 | (150,154) | 20 | (8,26) |
| **Baseline Dementia** |  |  |  |  |  |  |
| No Dementia | 129 | (124,136) | 111 | (110,113) | 18 | (13,23) |
| Dementia | 434 | (410,458) | 346 | (339,355) | 87 | (63,112) |

**Legend:** Standardized 90-day risk of mortality (events per 1000 individuals). CI, confidence intervals.

## DISCUSSION

Our results provide significant insights into the mortality risks associated with BZD initiation following AIS among Medicare beneficiaries. There was an excess 90-day mortality risk for patients who initiated BZDs within 30 days post-AIS discharge. This finding suggests that BZD use after an AIS among patients over 76 years may be associated with increased mortality risk. When we further analyzed by age groups, the mortality risk increased significantly with age; patients 86 years and older had a higher risk than the 76-80 group.

Additionally, more female adults were initiated on BZDs than male adults. Perhaps the most striking finding was the significant excess mortality among patients with baseline dementia. Therefore, initiating BZDs should be based on carefully evaluating individual patient characteristics and weighing the potential benefits against the increased mortality risk.

While anxiety or insomnia can be managed with BZDs in the short term, the potential risks and side effects are significant among older adults due to age-related changes in drug metabolism, polypharmacy, and comorbidities.^12,29^ Of concern, BZDs can impair cognition (i.e., excessive somnolence, drowsiness, dizziness, confusion, memory problems, and difficulties with attention, etc.), cause or aggravate often related psychomotor issues (i.e., gait and balance issues), and can lead to hip fracture, pneumonia, respiratory depression requiring intubation, and death.^5–11,30,31^ Lastly, the sedating effects of BZDs on ambulation dexterity might have a more significant impact among AIS survivors who can walk during the early days of recovery (e.g., those with mild AIS) and are at risk for falls and fall-related injuries.^6,10^

Despite state policies and national guidelines to curb BZD prescriptions, usage remains high,^4,14^ especially among older adults who often receive 180 doses or more.^32^ The prevalence of BZD usage among older adults in the United States increased from 1.1% to 17.6% between 2012 and 2013 (when BZDs were added to the Medicare formulary), with a BZD use rate ranging from 8.6% to 20% among adults 65 years and older.^33^ Of concern, many stroke patients receive BZD prescriptions indefinitely, and the majority using BZDs chronically experience adverse effects.^3,5–9,14,15^ While BZD initiation typically occurs at the hospital (where this type of medication might be of greater need), 4.7% of AIS survivors received a new BZD prescription within the 90-day post-discharge window, and most were female patients.^34^ We examined outcomes associated with initiating BZDs at discharge when there is a great concern for this population, as prescriptions might be unnecessary, are not as closely supervised, and frequency is not controlled. Long-term use of BZDs can lead to physical dependence, even at therapeutic doses. For older stroke survivors, this poses additional risks because sudden discontinuation can lead to withdrawal symptoms, including rebound anxiety, insomnia, depression, seizures, or delirium.^11,35,36^

There is scarce data and studies on older adult patient outcomes among those receiving BZD prescriptions. When trials are infeasible, comparative effectiveness and safety studies leveraging real-world practice variation offer a powerful alternative.^25,37,38^ Additionally, we used techniques developed by Hernan et al. that allowed us to address two primary sources of bias arising from structuring initiation and using retrospective data using cloning.^25–27^ The first, known as immortal time bias, is due to a differing discharge time and BZD initiation time, leading to spurious interpretations of this lag. The second, known as confounding bias due to randomization, arises in retrospective settings as we did not perform true randomization of our cohort. Using novel analytic strategies to emulate clinical trials that would otherwise be infeasible or unethical, our study proves the utility of these techniques. It presents meaningful results that should impact and reinforce BZD guidelines and improve health care for older adults.

Our study provides valuable evidence suggesting that BZD initiation after AIS is associated with increased mortality risk in Medicare beneficiaries, a greater risk observed predominantly among individuals 76 years and older, males, and those with dementia. These findings underscore the importance of careful consideration and individualized decision-making when managing post-stroke care in older adults, weighing risks against potential benefits, and considering alternative treatments when possible.

### Limitations

The statistical analysis is contingent on modeling the survival function across the 90-day observation period. Due to the large sample size and long observation window, statistical results are sensitive to the initial specifications of the outcome model. In Technical Details (Supplement), we provide further details about our outcome model’s exact specifications and justifications, which incorporate medical information and statistical parsimony.

The findings for our study population may not generalize to different age groups and patients with varying diagnoses or subtype etiology (i.e., intracerebral hemorrhage, infarction, cardiac embolism, etc.) and might not apply to Medicare Advantage beneficiaries. Due to data availability, our sample was limited to Traditional Medicare.

## CONCLUSION

Initiating BZDs within 30 days post-AIS discharge significantly increased the 90-day mortality risk among Medicare beneficiaries aged 76 and older and those with baseline dementia.

## Data Availability

We had a Data Use Agreement approved by the Center for Medicare & Medicaid Services (DUA RSCH-2022-58182). Interested researchers may replicate the study by obtaining the data from CMS. Reproducing this study requires 20% MedPAR, Outpatient, Carrier, and Part D standard analytical files.

## SUPPLEMENTAL MATERIALS

Tables S1–S4

Figure S1–S12

Technical Details

Statistical Code

## REFERENCES

1. 1. CDC. Stroke Facts [Internet]. Stroke. 2024 [cited 2024 Jul 10];Available from: https://www.cdc.gov/stroke/data-research/facts-stats/index.html

2. Ovbiagele B, Goldstein LB, Higashida RT, Howard VJ, Johnston SC, Khavjou OA, Lackland DT, Lichtman JH, Mohl S, Sacco RL, et al. Forecasting the future of stroke in the United States: a policy statement from the American Heart Association and American Stroke Association. Stroke. 2013;44:2361–2375.

3. Moura LMVR, Yan Z, Donahue MA, Smith LH, Schwamm LH, Hsu J, Newhouse JP, Haneuse S, Blacker D, Hernandez-Diaz S. No short-term mortality from benzodiazepine use post-acute ischemic stroke after accounting for bias. J Clin Epidemiol. 2023;154:136–145.

4. By the 2023 American Geriatrics Society Beers Criteria® Update Expert Panel. American Geriatrics Society 2023 updated AGS Beers Criteria® for potentially inappropriate medication use in older adults. J Am Geriatr Soc. 2023;71:2052–2081.

5. Lee JS. Necessary evil or systemic failure of care? Use of benzodiazepine and antipsychotic medications in older people seeking emergency department care. J Am Geriatr Soc. 2022;70:698– 700.

6. Díaz-Gutiérrez MJ, Martínez-Cengotitabengoa M, Sáez de Adana E, Cano AI, Martínez-Cengotitabengoa MT, Besga A, Segarra R, González-Pinto A. Relationship between the use of benzodiazepines and falls in older adults: A systematic review. Maturitas. 2017;101:17–22.

7. Vozoris NT, Fischer HD, Wang X, Stephenson AL, Gershon AS, Gruneir A, Austin PC, Anderson GM, Bell CM, Gill SS, et al. Benzodiazepine drug use and adverse respiratory outcomes among older adults with COPD. Eur Respir J. 2014;44:332–340.

8. Markota M, Rummans TA, Bostwick JM, Lapid MI. Benzodiazepine Use in Older Adults: Dangers, Management, and Alternative Therapies. Mayo Clin Proc. 2016;91:1632–1639.

9. Gupta A, Bhattacharya G, Balaram K, Tampi D, Tampi RR. Benzodiazepine use among older adults. Neurodegener Dis Manag. 2021;11:5–8.

10. Sun S, Lomachinsky V, Smith LH, Newhouse JP, Westover MB, Blacker D, Schwamm L, Haneuse S, Moura LMVR. Benzodiazepine Initiation and the Risk of Falls or Fall-Related Injuries in Older Adults Following Acute Ischemic Stroke. medRxiv. 2024;2024.02.06.24302430.

11. Olfson M, King M, Schoenbaum M. Benzodiazepine use in the United States. JAMA Psychiatry. 2015;72:136–142.

12. Pesante-Pinto JL. Clinical Pharmacology and the Risks of Polypharmacy in the Geriatric Patient. Phys Med Rehabil Clin N Am. 2017;28:739–746.

13. Lin S-M, Yang S-H, Liang C-C, Huang H-K, Loh C-H. Association between benzodiazepine use and risks of chronic-onset poststroke pneumonia: a population-based cohort study. BMJ Open. 2019;9:e024180.

14. Wang F, Ma Z, Liu M, Wu X. Potentially inappropriate medications at admission and discharge in older adults: A comparison of the Beers 2019 and 2015 criteria. Int J Clin Pharmacol Ther. 2020;58:299–309.

15. Agarwal SD, Landon BE. Patterns in Outpatient Benzodiazepine Prescribing in the United States. JAMA Netw Open. 2019;2:e187399.

16. Buttgereit T, Palmowski A, Forsat N, Boers M, Witham MD, Rodondi N, Moutzouri E, Navidad AJQ, Van’t Hof AWJ, van der Worp B, et al. Barriers and potential solutions in the recruitment and retention of older patients in clinical trials-lessons learned from six large multicentre randomized controlled trials. Age Ageing. 2021;50:1988–1996.

17. von Elm E, Altman DG, Egger M, Pocock SJ, Gøtzsche PC, Vandenbroucke JP, STROBE Initiative. Strengthening the Reporting of Observational Studies in Epidemiology (STROBE) statement: guidelines for reporting observational studies. BMJ. 2007;335:806–808.

18. Eicheldinger C, Bonito A. More accurate racial and ethnic codes for Medicare administrative data. Health Care Financ Rev. 2008;29:27–42.

19. McCormick N, Bhole V, Lacaille D, Avina-Zubieta JA. Validity of Diagnostic Codes for Acute Stroke in Administrative Databases: A Systematic Review. PLoS One. 2015;10:e0135834.

20. Runde D. Calculated Decisions: NIH stroke scale/score (NIHSS). Emerg Med Pract. 2020;22:CD6– CD7.

21. Schlegel D, Kolb SJ, Luciano JM, Tovar JM, Cucchiara BL, Liebeskind DS, Kasner SE. Utility of the NIH Stroke Scale as a predictor of hospital disposition. Stroke. 2003;34:134–137.

22. Han TS, Murray P, Robin J, Wilkinson P, Fluck D, Fry CH. Evaluation of the association of length of stay in hospital and outcomes. Int J Qual Health Care. 2021;34:mzab160.

23. Sud M, Yu B, Wijeysundera HC, Austin PC, Ko DT, Braga J, Cram P, Spertus JA, Domanski M, Lee DS. Associations Between Short or Long Length of Stay and 30-Day Readmission and Mortality in Hospitalized Patients With Heart Failure. JACC Heart Fail. 2017;5:578–588.

24. Moura LMVR, Festa N, Price M, Volya M, Benson NM, Zafar S, Weiss M, Blacker D, Normand S-L, Newhouse JP, et al. Identifying Medicare beneficiaries with dementia. J Am Geriatr Soc. 2021;69:2240–2251.

25. Hernán MA, Wang W, Leaf DE. Target Trial Emulation: A Framework for Causal Inference From Observational Data. JAMA. 2022;328:2446–2447.

26. Hernán MA, Robins JM. Using Big Data to Emulate a Target Trial When a Randomized Trial Is Not Available. Am J Epidemiol. 2016;183:758–764.

27. Hernán MA, Robins JM. Estimating causal effects from epidemiological data. J Epidemiol Community Health. 2006;60:578–586.

28. Wu C-C, Liao M-H, Su C-H, Poly TN, Lin M-C. Benzodiazepine Use and the Risk of Dementia in the Elderly Population: An Umbrella Review of Meta-Analyses. J Pers Med. 2023;13:1485.

29. Hoel RW, Giddings Connolly RM, Takahashi PY. Polypharmacy Management in Older Patients. Mayo Clin Proc. 2021;96:242–256.

30. Votaw VR, Geyer R, Rieselbach MM, McHugh RK. The epidemiology of benzodiazepine misuse: A systematic review. Drug Alcohol Depend. 2019;200:95–114.

31. Dublin S, Walker RL, Jackson ML, Nelson JC, Weiss NS, Von Korff M, Jackson LA. Use of opioids or benzodiazepines and risk of pneumonia in older adults: a population-based case-control study. J Am Geriatr Soc. 2011;59:1899–1907.

32. Shorr RI, Bauwens SF, Landefeld CS. Failure to limit quantities of benzodiazepine hypnotic drugs for outpatients: placing the elderly at risk. Am J Med. 1990;89:725–732.

33. Maust DT, Kim HM, Wiechers IR, Ignacio RV, Bohnert ASB, Blow FC. Benzodiazepine Use among Medicare, Commercially Insured, and Veteran Older Adults, 2013-2017. J Am Geriatr Soc. 2021;69:98–105.

34. Torres VL, Brooks JD, Donahue MA, Sun S, Hsu J, Price M, Blacker D, Schwamm L, Newhouse JP, Haneuse S, et al. Benzodiazepine Utilization in Ischemic Stroke Survivors: Analyzing Initial Excess Supply and Longitudinal Trends. Stroke. 2024; In press.

35. Juergens SM. Problems with benzodiazepines in elderly patients. Mayo Clin Proc. 1993;68:818– 820.

36. Maust DT, Petzold K, Strominger J, Kim HM, Bohnert ASB. Benzodiazepine Discontinuation and Mortality Among Patients Receiving Long-Term Benzodiazepine Therapy. JAMA Netw Open. 2023;6:e2348557.

37. Moura LMVR, Donahue MA, Yan Z, Smith LH, Hsu J, Newhouse JP, Schwamm LH, Haneuse S, Hernandez-Diaz S, Blacker D. Comparative Effectiveness and Safety of Seizure Prophylaxis Among Adults After Acute Ischemic Stroke. Stroke. 2023;54:527–536.

38. Moura LM, Westover MB, Kwasnik D, Cole AJ, Hsu J. Causal inference as an emerging statistical approach in neurology: an example for epilepsy in the elderly. Clin Epidemiol. 2017;9:9–18.

